# Manual Validation of an Algorithm for Identifying Osteoarthritis within the Centralized Interactive Phenomics Resource of the Million Veteran Program

**DOI:** 10.1101/2025.05.12.25327449

**Authors:** Chelsea T. Nguyen, Joshua S. Richman, Joe W. Chiles, Jasvinder A. Singh, Merry-Lynn N. McDonald

## Abstract

**Objective:** This study aimed to validate an osteoarthritis (OA) phenotyping algorithm within the Million Veteran Program (MVP) using the United States (US) Department of Veterans Affairs (VA) Centralized Interactive Phenomics Resource (CIPHER).

**Methods:** A random sample of 213 veterans was analyzed sing ICD-9-CM/ICD-10-CM codes from a previously published algorithm (PMID:29559693). OA cases required two OA codes at least 30 days apart, while controls were excluded based on codes for conditions more common in OA patients, such as chondrocalcinosis and crystal arthropathies. Manual chart reviews identified documented OA mentions and joint replacements. Cohen’s kappa statistic assessed agreement. Discrepancies between chart data and coding were re-evaluated through re-abstraction.

**Results:** Among 213 veterans, 174 (82%) had chart-documented OA. Agreement between chart review and code-based general OA identification was moderate (kappa = 0.47). Joint-specific agreement was substantial for knee (kappa = 0.63) and hip OA (kappa = 0.59), but lower for spine (kappa = 0.16) and hand (kappa = 0.34). Agreement was high for hip (kappa = 0.86) and knee replacements (kappa = 0.69). The McNemar test showed significant asymmetry for general OA, hand OA, and thumb OA, indicating discrepancies between chart and coded data. No significant asymmetry was found for knee and hip OA, supporting better alignment.

**Conclusions:** This study supports the validity of the OA phenotyping algorithm using the VA database for identifying OA. The variability in identifying milder cases highlights the need for refined phenotyping algorithms and standardized diagnostic protocols to improve OA detection and personalized care for veterans.

## INTRODUCTION

Osteoarthritis (OA) is a major public health concern, with an estimated 32.5 million adults in the United States (US) affected by this debilitating disease. The societal and patient-centered impacts of end-stage or advanced OA are particularly significant among United States veterans^1^. Veterans with advanced OA frequently undergo total hip arthroplasty (THA) or total knee arthroplasty (TKA)^2^ and subsequent rehabilitation, yet face diminished health-related quality of life (HRQL)^3^, persistent functional limitations, and a higher incidence of comorbidities, hospitalization, and health care access^4^. Despite the high burden of OA, its underlying etiology remains poorly understood, limiting the development of effective treatments. These efforts are hampered by the need for reliable OA phenotyping algorithms to harness critical information in large hospital and population biobanks,

The Veterans Affairs (VA) is the largest integrated healthcare system in the US, serving over five million veterans annually^5^. Our research and that of others have identified genetic risk variants associated with OA^6^ using self-reported disease status^7^ and ICD code-based definitions^8,9^. Analysis of Electronic Medical Record (EMR) data can yield insights into potential therapeutic treatment interventions, complementing other data sources and facilitating larger sample sizes^10^. However, results can be prone to biases and errors arising from data collection intended primarily for clinical care and billing purposes^11^. Errors may stem from inaccurate patient observations and recording of results, necessitating extensive quality control, phenotyping, and extraction strategies^11^.

Despite the critical need for accurate OA phenotyping, there is a notable gap in the literature. Few validation studies^12-17^ have systematically assessed the accuracy of OA diagnoses within EMR databases. To enhance validity, some studies have incorporated stricter criteria, such as requiring at least one diagnostic code in conjunction with documentation of OA-related treatment (e.g., prescribed medications, physical therapy, or surgical intervention)^18^ or two diagnostic codes recorded at separate time points to confirm a persistent diagnosis^9,19^. These methods improve specificity by reducing the likelihood of capturing incidental or incorrect OA diagnoses. Accurate phenotyping is essential for several reasons: it allows for personalized treatment approaches tailored to veterans, enhancing the likelihood of successful outcomes by predicting treatment responses based on specific phenotypes. Additionally, it improves prognosis and disease management by providing insights into disease progression and identifying comorbidities associated with different OA types. This understanding can lead to more proactive management strategies and better integrated care. Moreover, enhanced research efforts benefit from accurate phenotyping by standardizing clinical trial outcomes and facilitating the discovery of biomarkers linked to specific OA presentations. Finally, accurate phenotyping optimizes healthcare resource allocation, potentially reducing the economic burden of OA management. However, validation efforts remain understudied, and there is limited consensus on the optimal approach for defining OA cases within EMR databases.

Most validation studies have predominantly focused on knee OA^18^, with minimal exploration of other commonly affected joints such as the hip or spine. Given that OA is a heterogeneous disease with joint-specific variations in progression, symptom severity, and treatment response, a broader validation approach is necessary to capture the full spectrum of OA phenotypes. Addressing these limitations is crucial for ensuring the accuracy of EMR-based OA research, as well as for guiding clinical decision making and healthcare policy. Our study aims to address this gap by systemically validating our OA case and control definitions within the Million Veteran Program (MVP) dataset using comprehensive chart reviews conducted within the Centralized Interactive Phenomics Resource (CIPHER) database. This effort will provide a robust framework for future research, enabling more precise epidemiological studies, genetic analyses, and healthcare utilization assessments related to OA.

## METHODS

### IRB Approval

All participants were recruited as part of the MVP^20^. All participants had previously consented to sharing their deidentified data for research. The research described in this manuscript received ethical and study protocol approval from the Veterans Affairs Central Institutional Review Board in accordance with the principles outlined in the Declaration of Helsinki.

### CIPHER

The VA’s Centralized Interactive Phenomics Resource (CIPHER) serves as a cutting-edge knowledge base of computable EMR-based phenotypes derived from clinical health record system data, registries, and surveys^21^. CIPHER enhances the reproducibility, consistency, and scalability of phenotyping across health systems, offering a high-level overview of the development and validation process^21^.

### Procedure/Data Sources

This retrospective study employed an ICD (International Classification of Disease) algorithm to select a random sample of veterans with visits (outpatient, inpatient, or emergency care) for manual chart review, regardless of whether they had a documented diagnosis of OA (ICD-9/ICD-10 codes) at the Birmingham VA medical center.

### Sampling Methodology

A random sample of 213 veterans from a total of 354,628 veterans was selected based on phenotypic categories to evaluate concordance and discordance between ICD-9/ICD-10 diagnostic codes and free-text documentation in patient histories. Cases and controls were identified using ICD codes, and their medical records were meticulously abstracted and reviewed in a blinded process by Chelsea Nguyen and Dr. Joshua Richman. The sample was divided into five subsets: four subsets of 50 veterans each and one subset of 13 veterans. Additional chart review were conducted for hip and knee OA to assess decreased mobility, use of assistive devices, and joint replacement (hip, knee) mentions in clinical notes that were not captured by Current Procedural Terminology (CPT) codes for total joint replacement at non-VA facilities. CPT codes were utilized to identify and categorize relevant medical procedures, such as hip replacement and knee replacement. The total sample included only veterans classified as OA cases or controls based on the code-based definition, with an inclusive age range of 40 and 85 years of age.

### OA Case/Control Criteria

The criteria for OA case identification were defined using ICD-9/ICD-10 phenotype coding. A case was established as having at least two OA codes recorded at least 30 days apart, irrespective of category. A detailed list of codes was provided, encompassing general OA categories, and overlapping lists for specific subtypes (e.g., hip, knee, spine, hand, finger, and thumb) of OA (**Supplementary Table 1**). OA controls were excluded if they had codes for concomitant findings typically prevalent in osteoarthritis patients, including traumatic and post-traumatic arthropathy (**Supplementary Table 2**).

### ICD Codes

The relevant ICD-9-CM codes for OA includes ‘716.1, 716.10, 716.11, 716.12, 716.13, 716.14, 716.15, 716.16, 716.17, 716.18, 716.19’ and ICD-10-CM codes include ‘M12.50, M12.519, M12.529, M12.539, M12.549, M12.559, M12.569, M12.579, M12.58, M12.59, M19.1, M19.10, M19.11, M19.12, M19.13, M19.14, M19.15, M19.16, M19.17, M19.18, M19.19.’ Notably, codes ‘716.10, M12.50, M12.519, M12.529, M12.539, M12.549, M12.559, M12.569, M12.579, M12.58, M12.59’ were excluded from the control criteria.

### Validation of Chart Review Performance

Given the absence of an accepted gold standard for diagnosing OA, the study aimed to assess the inter-rater reliability or agreement between chart review findings and the code-based definition of OA. Two blinded reviewers independently evaluated VA EMRs for documentation of OA-related terms (e.g., Degenerative Joint Disease, Osteoarthritis, Arthritis) within key sections of patient records, including the chief complaint, history of present illness, and assessment and plan of each visit. Reviewers classified individuals based on chart documentation of general OA and OA at specific joint sites (hip, knee, spine, hand, and thumb), noting the first date of mention for each. Indications of hip or knee replacement were similarly reviewed and documented. Discrepancies between reviewers were adjudicated by a practicing rheumatologist, Dr. Jasvinder Singh, to ensure consistency and accuracy.

### Statistical Analyses

These statistics were calculated in R Studio (R version 4.3.1) using the Veterans Affairs Informatics and Computing Infrastructure (VINCI) platform. The percentage of individuals identified by code-based definitions within the charts, specifically examining the proportion of code-based controls whose chart documentation corroborated an OA diagnosis, was evaluated. To assess the agreement between chart review findings and the code-based definition, we calculated the kappa statistic, a measure of inter-rater reliability^22^. The kappa statistic is defined as: (observed agreement−expected agreement)/(1−expected agreement)^22^. Given the absence of a gold standard for OA diagnosis, the kappa statistic provides a framework for evaluating the consistency between these two approaches. Unweighted kappa statistics were calculated for each joint site and for joint replacement types to quantify agreement. As a secondary analysis, we employed the McNemar^23^ test for symmetry was employed to identify discrepancies between chart documentation and ICD code-based definitions, assessing whether the proportions of discordant cases (e.g., ICD code present but no chart mention or vice versa) were significantly different. This test was conducted with and without continuity correction, and p-values were used to evaluate statistical significance For discrepancies where chart documentation contradicted ICD code-based definitions, a re-abstraction process was performed. Chart abstraction findings were classified into three categories based on the presence of OA documentation. Definitive evidence referred to consistent documentation of OA, aligning with both ICD codes and chart reviews. Possible evidence indicated mentions of OA or related conditions (e.g., arthritis, degenerative joint disease) in the problem list but not as the primary diagnosis. No evidence was defined as the absence of OA documentation, with other conditions recorded as the primary diagnosis.

## RESULTS

### Demographics of patients included in analyses

A random sample of 213 veterans out of 354,628 veterans was included in the study, with OA mentions and ICD code-based classifications analyzed across different joint sites and joint replacement procedures. Cohort demographics, including age, sex, race, and period of military service, are summarized (**Table 1**). This analysis aimed to assess the concordance between OA mentions in patient charts and corresponding ICD code-based diagnoses, particularly differentiating between general OA (non-site-specific mentions), OA at specific joint sites (e.g., hip, knee, spine, hand, thumb), and joint replacements (e.g., hip, knee).

### Agreement between chart mentions and code-based definition in OA-associated joint sites

Among the 213 veterans, 82% had documented mentions of OA in their charts, while 18% did not (**Table 2**). When comparing chart mentions to the ICD-based definition, general OA showed moderate agreement with a kappa statistic of 0.48, suggesting discrepancies where OA was documented in the chart mentions but not coded specifically. The analysis also examined OA associated with specific joint sites, including the thumb, hand, hip, knee, and spine, as well as hip and knee replacements (**Table 2**). For hip OA, 56 patients (26%) had chart mentions, with a kappa statistic of 0.59, 85% observed agreement, and 63% expected agreement. Knee OA was mentioned in 89 cases (42%), with 65 identified as ICD-based cases, demonstrating substantial agreement (kappa = 0.63) with 82% observed agreement and 52% expected agreement. Spine OA had 67 chart mentions (31%) but showed poor agreement (kappa = 0.16), with 67% observed agreement and 60% expected agreement, indicating that spine-related diagnoses may be prioritized over OA in coding. Hand OA was recorded in 31 cases (15%), with 20 matching ICD codes (kappa = 0.34, fair agreement), while thumb OA had only 9 chart mentions (4%), with 3 corresponding ICD-based cases, showing slight agreement (kappa = 0.12) and suggesting that these sites may be underrepresented in ICD coding.

### Agreement between chart mentions and code-based definition in joint replacements

For joint replacements, documentation and coding aligned more closely. Hip replacement was mentioned in 25 cases (12%), with 22 having corresponding codes (kappa = 0.86, excellent agreement), with 97% observed agreement and 79% expected agreement (**Table 2**). Knee replacement had 29 mentions (14%), with 20 ICD-based cases (kappa = 0.69, substantial agreement), with 93% observed agreement and 78% expected agreement (**Table 2**). The strong agreement for joint replacements likely reflects the standardized documentation required for surgical procedures, making ICD coding more reliable.

### McNemar test analysis of symmetry in disagreement between chart documentation and ICD codes

The McNemar test assessed asymmetry in classification between chart documentation and ICD codes (**Table 2**). General OA showed significant asymmetry (χ^2^ = 14, p = 1.8E-04), indicating that chart documentation captured more cases than ICD codes, suggesting potential under-coding of OA cases. Hand OA (χ^2^ = 12, p = 6.9E-04) and thumb OA (χ^2^ = 8.04, p = 4.6E-03) also showed significant discrepancies, reinforcing the trend that milder OA cases are less frequently coded. Conversely, knee OA (χ^2^ = 2.1, p = 0.14) and hip OA (χ^2^ = 2.5, p = 0.11) did not exhibit significant asymmetry, indicating more consistent classification between chart mentions and ICD codes. Spine OA (χ^2^ = 5.6, p = 1.8E-02) displayed significant asymmetry, consistent with its low kappa statistic (0.16), further highlighting substantial coding discrepancies. In contrast, hip and knee replacements demonstrated no significant asymmetry (χ^2^ = 0, p = 1 for hip; χ^2^ = 0.27, p = 0.61 for knee), supporting their strong agreement across both documentation methods.

### Chart re-abstractions to address discrepancies between chart mentions and code-based definition

To further investigate discrepancies, a re-abstraction process was conducted for 23 instances where there was some evidence of OA in patient charts but no ICD codes. Of these 7 instances, we found strong evidence of OA in patient charts, while 14 instances (61%) were less conclusive where OA or related terms (e.g., Degenerative Joint Disease) were documented but did not seem to suggest an active or ongoing medical problem. In 2 instances (9%), we found no evidence of OA in patient charts. These findings suggest that ICD coding may underrepresent OA cases, particularly for milder presentations where OA is not the primary focus of any clinical visit or where symptoms are not severe enough to require treatment. This discrepancy may arise due to coding prioritization (where physicians focus on more clinically pressing conditions), variability in documentation styles (some providers record OA in free-text notes without assigning specific codes), or non-VA diagnoses (where veterans receive OA treatment outside the VA system, resulting in missing codes).

## DISCUSSION

This study validated the use of VA administrative data for the MVP OA phenotyping algorithm, demonstrating its potential for large-scale OA research. Moderate to substantial agreement was observed between chart documentation and our ICD code-based definition across various OA sites, though agreement varied by joint location. Hip replacement showed excellent agreement, followed by substantial agreement for knee replacement and knee OA. The re-abstraction process revealed that limiting analyses to cases with definitive chart evidence improved agreement metrics, underscoring the importance of stringent validation criteria to minimize bias. These findings highlight the need for enhanced diagnostic protocols and the integration of additional data sources—such as imaging studies, patient-reported outcomes, and longitudinal clinical data—to improve OA phenotyping accuracy.

Our findings align with previous studies using a gold standard^18^, requiring two diagnostic codes within 6 to 12 months significantly improved knee OA phenotyping accuracy. Yau *et al*. ^18^ conducted a knee OA validation study using a random sample from The Health Improvement Network (THIN) of patients aged 40-90 years, identifying cases based on their first diagnostic code for knee OA or knee pain. In this study, the gold standard was defined as knee pain with x-ray and/or MRI-confirmed OA. They reported high specificity using ICD codes for knee OA, indicating that code-based OA diagnoses were generally accurate, but low sensitivity, meaning many true OA cases were missed when replying on a single diagnostic code. This aligns with our findings, where hip and knee OA showed moderate to substantial concordance in VA administrative data.

Although, A key limitation of our study is the absence of a universally accepted gold standard for OA diagnosis within the VA system. While chart reviews provide valuable insights, they may overlook inconsistently documented cases, while ICD codes may fail to capture patients without formal diagnoses. Additionally, the Yau et al. ^18^ study focused solely on knee OA, limiting the generalizability of its findings to other joints. Despite these challenges, administrative codes for joint replacements proved to be reliable indicators of severe OA. This reliability likely stems from the structured nature of procedural coding, which is less prone to misclassification than diagnostic codes. Based on these findings, we recommend that EMR-based algorithms require at least two OA diagnostic codes to enhance accuracy.

On the other hand, milder OA cases (e.g., spine, hand, thumb OA) exhibited greater discrepancies, highlighting broader challenges in OA classification. Various studies have assessed the validity of using a single ICD-based diagnosis code in large datasets, including EMR and administrative data^15-17^. However, in our study spine, hand, and thumb OA showed particularly high discrepancies, reinforcing concerns that milder OA cases are systematically underrepresented due to inconsistent documentation and the absence of radiographic confirmation.

A key strength of our study is its comprehensive assessment of OA across multiple joint sites, including the hip, knee, spine, hand, and thumb, as well as joint replacement procedures. By incorporating both OA diagnoses and surgical interventions, we provided a more complete evaluation of how OA is documented in administrative data versus clinical records. The chart re-abstraction findings supported this approach, revealing that many code-based controls documented in clinical notes lacked corresponding ICD codes, often because OA was not listed as the primary diagnosis. Discrepancies between clinical documentation and ICD coding were particularly pronounced for spine, hand, and thumb OA, consistent with previous research showing that severe OA cases (e.g., those requiring joint replacement) are more reliably captured in administrative data, while milder cases often go undiagnosed or unrecorded. This has important implications for OA phenotyping, as relying solely on ICD codes may lead to under-detection of early-stage or mild OA, particularly in joints less commonly treated with surgical interventions. Given that early diagnosis and intervention can slow disease progression and improve patient outcomes, developing more sensitive phenotyping strategies is critical.

Despite these strengths, identifying milder OA cases remains a significant challenge, particularly those presenting with intermittent pain, stiffness, or without joint replacement. Clinicians may document OA in free-text notes without assigning a corresponding ICD code, and coding prioritization may focus on more pressing conditions, leading to underrepresentation of OA. Our chart re-abstraction analysis provided deeper insights into these discrepancies. Milder OA cases are less likely to undergo imaging, and veterans receiving care outside the VA system may have missing codes in VA records, further reducing standardization in documentation.

In conclusion, this study demonstrates the value of VA administrative data as a reliable resource for identifying OA-related healthcare utilization, while emphasizing the need for rigorous validation to improve phenotyping precision. Requiring at least two OA codes 30 days apart or greater improves the MVP algorithm’s reliability and validity, particularly for identifying severe cases of OA. The complementary use of kappa coefficients and McNemar’s test provides a robust framework for evaluating agreement and identifying discrepancies. However, further refinement is needed to capture early and milder cases, particularly in joint sites that are not as common. Addressing these inconsistencies through refining diagnostic algorithms and multimodal data integration will enhance OA phenotyping accuracy, leading to improved disease understanding, treatment outcomes, and healthcare delivery. Further research, including larger and more diverse cohorts, is needed to optimize diagnostic algorithms and ensure their reliability in identifying OA across different joint sites and stages of disease. Accurate OA phenotyping is critical for tailoring treatment strategies and improving healthcare delivery. Misclassification can have significant clinical implications, particularly for veterans who may benefit from early interventions to prevent disease progression. Implementing standardized diagnostic protocols and enhanced validation processes within EMR systems could reduce errors, improve consistency in OA identification, and enhance patient outcomes, ultimately lowering healthcare costs.

## Data Availability

This study relied on the electronic health records housed in the VA medical system. All summary-level data produced in the present study are available upon reasonable request to the authors

## AUTHOR CONTRIBUTIONS

Conception and design of research: JAS, MLM; Performed experiments: CN, JR, JC; Drafted manuscript: CN; Prepared figures and tables: JR, CN; Provided feedback on first draft of manuscript: JAS, MLM, JC; Approved final version of manuscript: MLM, JAS, JC, JR, CN

## FUNDING

This research is based on data from the Million Veteran Program, Office of Research and Development, Veterans Health Administration, and was supported by MVP000, MVP078 as well as award #I01RX002745. This publication does not represent the views of the Department of Veteran Affairs or the United States Government.

## DISCLOSURES

JAS has received consultant fees from ROMTech, Atheneum, Clearview healthcare partners, American College of Rheumatology, Yale, Hulio, Horizon Pharmaceuticals/DINORA, ANI/Exeltis, USA Inc., Frictionless Solutions, Schipher, Crealta/Horizon, Medisys, Fidia, PK Med, Two labs Inc., Adept Field Solutions, Clinical Care options, Putnam associates, Focus forward, Navigant consulting, Spherix, MedIQ, Jupiter Life Science, UBM LLC, Trio Health, Medscape, WebMD, and Practice Point communications; the National Institutes of Health; and the American College of Rheumatology. JAS has received institutional research support from Zimmer Biomet Holdings. JAS received food and beverage payments from Intuitive Surgical Inc./Philips Electronics North America. JAS owns stock options in Atai life sciences, Kintara therapeutics, Intelligent Biosolutions, Acumen pharmaceutical, TPT Global Tech, Vaxart pharmaceuticals, Atyu biopharma, Adaptimmune Therapeutics, GeoVax Labs, Pieris Pharmaceuticals, Enzolytics Inc., Seres Therapeutics, Tonix Pharmaceuticals Holding Corp., Aebona Pharmaceuticals, and Charlotte’s Web Holdings, Inc. JAS previously owned stock options in Amarin, Viking and Moderna pharmaceuticals. JAS is on the speaker’s bureau of Simply Speaking. Dr. Singh was a member of the executive of Outcomes Measures in Rheumatology (OMERACT), an organization that develops outcome measures in rheumatology and receives arms-length funding from 8 companies. Dr. Singh serves on the FDA Arthritis Advisory Committee. Dr. Singh is the co-chair of the Veterans Affairs Rheumatology Field Advisory Board (FAB). Dr. Singh is the editor and the Director of the University of Alabama at Birmingham (UAB) Cochrane Musculoskeletal Group Satellite Center on Network Meta-analysis. Dr. Singh previously served as a member of the following committees: member, the American College of Rheumatology’s (ACR) Annual Meeting Planning Committee (AMPC) and Quality of Care Committees, the Chair of the ACR Meet-the-Professor, Workshop and Study Group Subcommittee and the co-chair of the ACR Criteria and Response Criteria subcommittee. Other authors have no conflict of interest to disclose.

## ACKNOWLEDGEMENTS

The authors thank the Million Veteran Program (MVP) staff, researchers, and volunteers, who have contributed to MVP, and especially who previously served their country in the military and now generously agreed to enroll in the study (see mvp.va.gov for more information). The underlying work was based on data from the Million Veteran Program, Office of Research and Development, and the Veterans Health Administration.

## VA Million Veteran Program: Core Acknowledgement

### MVP Program Office

- Sumitra Muralidhar, Ph.D., Program Director US Department of Veterans Affairs, 810 Vermont Avenue NW, Washington, DC 20420
- Jennifer Moser, Ph.D., Associate Director, Scientific Programs US Department of Veterans Affairs, 810 Vermont Avenue NW, Washington, DC 20420
- Jennifer E. Deen, B.S., Associate Director, Cohort & Public Relations US Department of Veterans Affairs, 810 Vermont Avenue NW, Washington, DC 20420

### MVP Executive Committee

- Co-Chair: Philip S. Tsao, Ph.D. VA Palo Alto Health Care System, 3801 Miranda Avenue, Palo Alto, CA 94304
- Co-Chair: Sumitra Muralidhar, Ph.D. US Department of Veterans Affairs, 810 Vermont Avenue NW, Washington, DC 20420
- J. Michael Gaziano, M.D., M.P.H. VA Boston Healthcare System, 150 S. Huntington Avenue, Boston, MA 02130
- Elizabeth Hauser, Ph.D. Durham VA Medical Center, 508 Fulton Street, Durham, NC 27705
- Amy Kilbourne, Ph.D., M.P.H. VA HSR&D, 2215 Fuller Road, Ann Arbor, MI 48105
- Michael Matheny, M.D., M.S., M.P.H. VA Tennessee Valley Healthcare System, 1310 24th Ave. South, Nashville, TN 37212
- Dave Oslin, M.D. Philadelphia VA Medical Center, 3900 Woodland Avenue, Philadelphia, PA 19104
- Deepak Voora, MD Durham VA Medical Center, 508 Fulton Street, Durham, NC 27705

### MVP Co-Principal Investigators

- J. Michael Gaziano, M.D., M.P.H. VA Boston Healthcare System, 150 S. Huntington Avenue, Boston, MA 02130
- Philip S. Tsao, Ph.D. VA Palo Alto Health Care System, 3801 Miranda Avenue, Palo Alto, CA 94304

### MVP Core Operations

- Jessica V. Brewer, M.P.H., Director, MVP Cohort Operations VA Boston Healthcare System, 150 S. Huntington Avenue, Boston, MA 02130
- Mary T. Brophy M.D., M.P.H., Director, VA Central Biorepository VA Boston Healthcare System, 150 S. Huntington Avenue, Boston, MA 02130
- Kelly Cho, M.P.H, Ph.D., Director, MVP Phenomics VA Boston Healthcare System, 150 S. Huntington Avenue, Boston, MA 02130
- Lori Churby, B.S., Director, MVP Regulatory Affairs VA Palo Alto Health Care System, 3801 Miranda Avenue, Palo Alto, CA 94304
- Scott L. DuVall, Ph.D., Director, VA Informatics and Computing Infrastructure (VINCI) VA Salt Lake City Health Care System, 500 Foothill Drive, Salt Lake City, UT 84148
- Saiju Pyarajan Ph.D., Director, Data and Computational Sciences VA Boston Healthcare System, 150 S. Huntington Avenue, Boston, MA 02130
- Robert Ringer, Pharm.D., Director, VA Albuquerque Central Biorepository New Mexico VA Health Care System, 1501 San Pedro Drive SE, Albuquerque, NM 87108
- Luis E. Selva, Ph.D., Director, MVP Biorepository Coordination VA Boston Healthcare System, 150 S. Huntington Avenue, Boston, MA 02130
- Shahpoor (Alex) Shayan, M.S., Director, MVP PRE Informatics VA Boston Healthcare System, 150 S. Huntington Avenue, Boston, MA 02130
- Brady Stephens, M.S., Principal Investigator, MVP Information Center Canandaigua VA Medical Center, 400 Fort Hill Avenue, Canandaigua, NY 14424
- Stacey B. Whitbourne, Ph.D., Director, MVP Cohort Development and Management VA Boston Healthcare System, 150 S. Huntington Avenue, Boston, MA 02130

### MVP Publications and Presentations Committee

- Co-Chair: Themistocles L. Assimes, M.D., Ph. D VA Palo Alto Health Care System, 3801 Miranda Avenue, Palo Alto, CA 94304
- Co-Chair: Adriana Hung, M.D.; M.P.H VA Tennessee Valley Healthcare System, 1310 24^th^ Ave. South, Nashville, TN 37212
- Co-Chair: Henry Kranzler, M.D. Philadelphia VA Medical Center, 3900 Woodland Avenue, Philadelphia, PA 19104

**Table 1:**
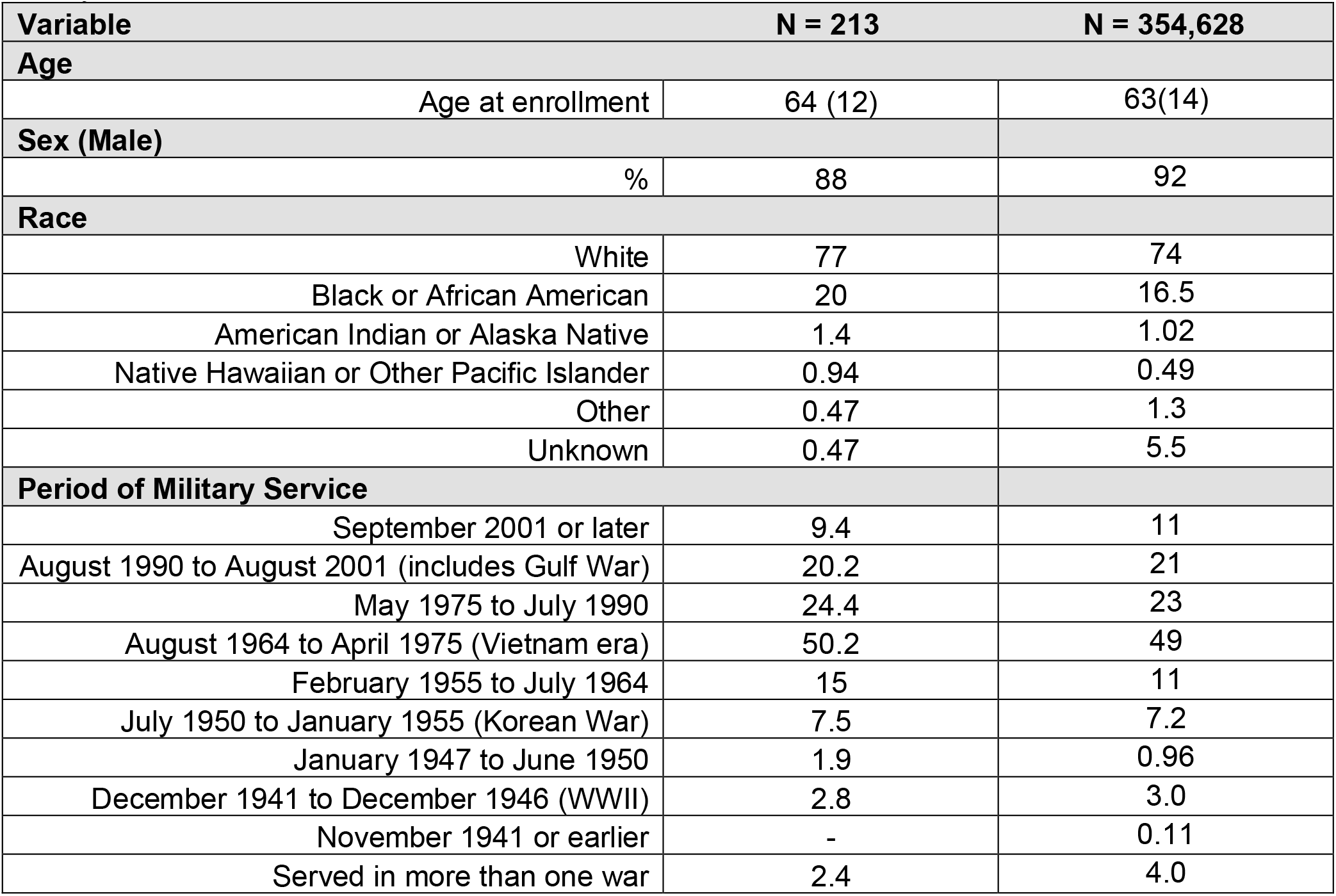
Demographics of a random sample of 213 Veterans out of 354,628 Veterans in the Million Veteran Program included in this validation study. The total sample size of 354,628 Veterans only included those who were identified as an OA case or control by the code-based definition. Values are reported as mean (standard deviation) for age and percentage (%) for male sex, race, and period of military service. OA, Osteoarthritis.

**Table 2:**
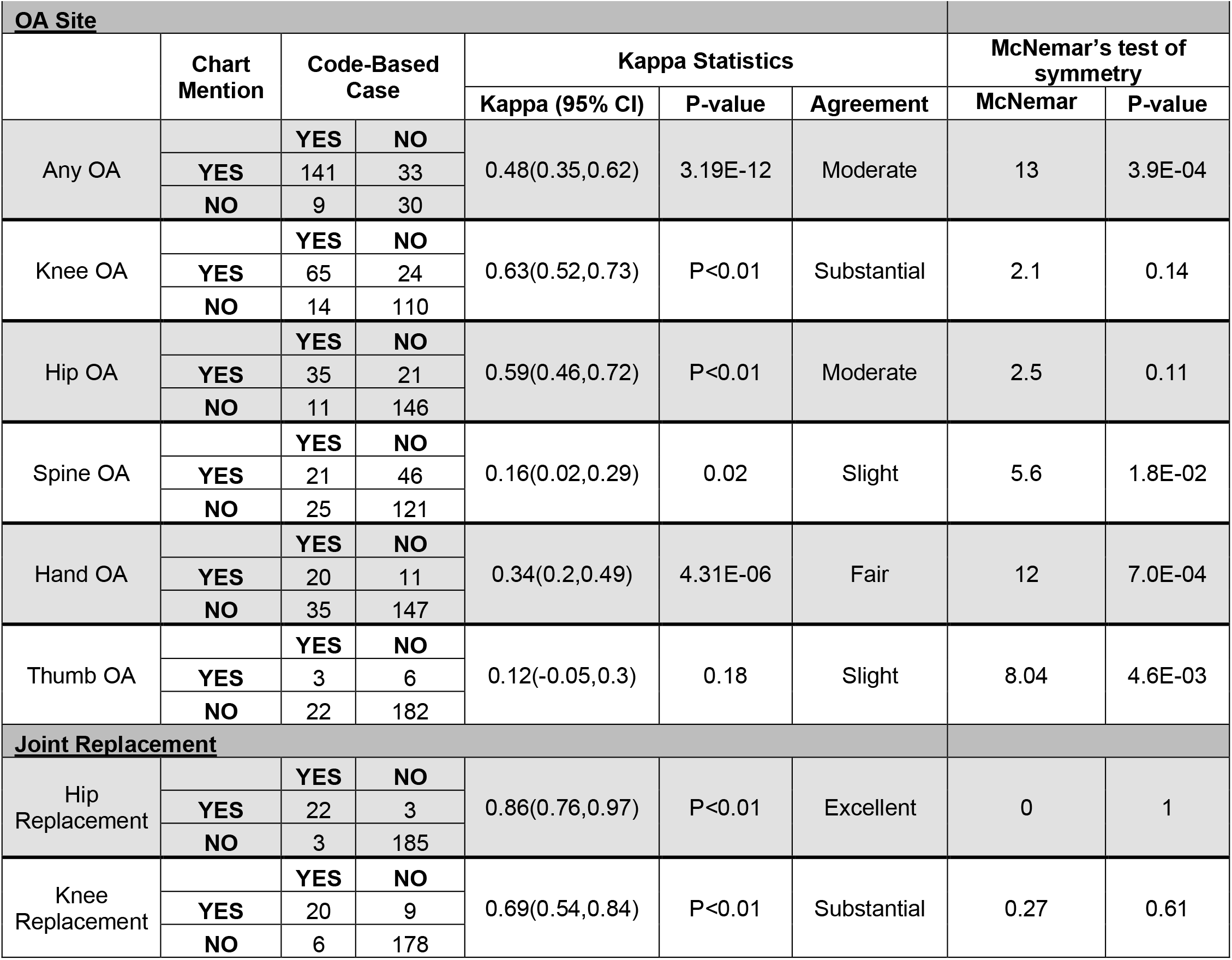
Concordance between chart mentions and ICD-code-based definition in a random sample of 213 Million Veteran Program Veterans with chart abstraction based on OA site and joint replacement. Values represent the concordance between chart-documented OA mentions and ICD-code-based definitions for each OA site and joint replacement. Kappa values indicate the level of agreement: <0.20 = slight, 0.21–0.40 = fair, 0.41–0.60 = moderate, 0.61–0.80 = substantial, and >0.81 = excellent. McNemar test results of symmetry in discordant classifications between chart mentions and ICD-code-based definition of OA. OA, Osteoarthritis; 95% CI, 95% confidence interval. McNemar’s chi-square (χ2) values are reported with continuity correction. A significant McNemar test (*p* < 0.05) indicates asymmetry in discordant classifications, meaning one method systematically identified more cases than the other.

| OA Site |  |  |  |  |  |  |  |  |
| --- | --- | --- | --- | --- | --- | --- | --- | --- |
|  | Chart Mention | Code-Based Case |  | Kappa Statistics |  |  | McNemar's test of symmetry |  |
|  |  |  |  | Kappa (95% CI) | P-value | Agreement | McNemar | P-value |
| Any OA |  | YES | NO | 0.48(0.35,0.62) | 3.19E-12 | Moderate | 13 | 3.9E-04 |
|  | YES | 141 | 33 |  |  |  |  |  |
|  | NO | 9 | 30 |  |  |  |  |  |
| Knee OA |  | YES | NO | 0.63(0.52,0.73) | P<0.01 | Substantial | 2.1 | 0.14 |
|  | YES | 65 | 24 |  |  |  |  |  |
|  | NO | 14 | 110 |  |  |  |  |  |
| Hip OA |  | YES | NO | 0.59(0.46,0.72) | P<0.01 | Moderate | 2.5 | 0.11 |
|  | YES | 35 | 21 |  |  |  |  |  |
|  | NO | 11 | 146 |  |  |  |  |  |
| Spine OA |  | YES | NO | 0.16(0.02,0.29) | 0.02 | Slight | 5.6 | 1.8E-02 |
|  | YES | 21 | 46 |  |  |  |  |  |
|  | NO | 25 | 121 |  |  |  |  |  |
| Hand OA |  | YES | NO | 0.34(0.2,0.49) | 4.31E-06 | Fair | 12 | 7.0E-04 |
|  | YES | 20 | 11 |  |  |  |  |  |
|  | NO | 35 | 147 |  |  |  |  |  |
| Thumb OA |  | YES | NO | 0.12(-0.05,0.3) | 0.18 | Slight | 8.04 | 4.6E-03 |
|  | YES | 3 | 6 |  |  |  |  |  |
|  | NO | 22 | 182 |  |  |  |  |  |
| Joint Replacement |  |  |  |  |  |  |  |  |
| Hip Replacement |  | YES | NO | 0.86(0.76,0.97) | P<0.01 | Excellent | 0 | 1 |
|  | YES | 22 | 3 |  |  |  |  |  |
|  | NO | 3 | 185 |  |  |  |  |  |
| Knee Replacement |  | YES | NO | 0.69(0.54,0.84) | P<0.01 | Substantial | 0.27 | 0.61 |
|  | YES | 20 | 9 |  |  |  |  |  |
|  | NO | 6 | 178 |  |  |  |  |  |

## Notes

### Author Declarations

All participants were recruited as part of the MVP. All participants had previously consented to sharing their deidentified data for research. The research described in this manuscript received ethical and study protocol approval from the Veterans Affairs Central Institutional Review Board in accordance with the principles outlined in the Declaration of Helsinki.

## REFERENCES

1. Singh, J.A. et al. Mediation of smoking-associated postoperative mortality by perioperative complications in veterans undergoing elective surgery: data from Veterans Affairs Surgical Quality Improvement Program (VASQIP)—a cohort study. BMJ Open 3(2013).

2. Singh, J.A. & Sloan, J.A. Health-related quality of life in veterans with prevalent total knee arthroplasty and total hip arthroplasty. Rheumatology (Oxford) 47, 1826–31 (2008).

3. Singh, J.A. & Murdoch, M. Effect of health-related quality of life on women and men’s Veterans Affairs (VA) health care utilization and mortality. J Gen Intern Med 22, 1260–7 (2007).

4. Singh, J.A., Nelson, D.B., Fink, H.A. & Nichol, K.L. Health-related quality of life predicts future health care utilization and mortality in veterans with self-reported physician-diagnosed arthritis: the veterans arthritis quality of life study. Semin Arthritis Rheum 34, 755–65 (2005).

5. Perlin, J.B., Kolodner, R. M., & Roswell, R. H. The Veterans Health Administration: quality, value, accountability, and information as transforming strategies for patient-centered care. The American journal of managed care 10, 828–836 (2004).

6. McDonald, M.-L.N. et al. Novel genetic loci associated with osteoarthritis in multi-ancestry analyses in the Million Veteran Program and UK Biobank. Nature Genetics 54, 1816–1826 (2022).

7. Zengini, E. et al. Genome-wide analyses using UK Biobank data provide insights into the genetic architecture of osteoarthritis. Nat Genet 50, 549–558 (2018).

8. Boer, C.G. et al. Deciphering osteoarthritis genetics across 826,690 individuals from 9 populations. Cell 184, 4784–4818 e17 (2021).

9. McDonald, M.N. et al. Novel genetic loci associated with osteoarthritis in multi-ancestry analyses in the Million Veteran Program and UK Biobank. Nat Genet 54, 1816–1826 (2022).

10. Katkade, V.B., Sanders, K.N. & Zou, K.H. Real world data: an opportunity to supplement existing evidence for the use of long-established medicines in health care decision making. J Multidiscip Healthc 11, 295–304 (2018).

11. Young, J.C., Conover, M.M. & Funk, M.J. Measurement error and misclassification in electronic medical records: methods to mitigate bias. Curr Epidemiol Rep 5, 343–356 (2018).

12. Singh, J.A. Veterans Affairs databases are accurate for gout-related health care utilization: a validation study. Arthritis research & therapy 15, R224 (2013).

13. Rahman, M.M., Kopec, J.A., Goldsmith, C.H., Anis, A.H. & Cibere, J. Validation of Administrative Osteoarthritis Diagnosis Using a Clinical and Radiological Population-Based Cohort. Int J Rheumatol 2016, 6475318 (2016).

14. Turkiewicz, A. et al. Current and future impact of osteoarthritis on health care: a population-based study with projections to year 2032. Osteoarthritis Cartilage 22, 1826–32 (2014).

15. Funck-Brentano, T., Nethander, M., Moverare-Skrtic, S., Richette, P. & Ohlsson, C. Causal Factors for Knee, Hip, and Hand Osteoarthritis: A Mendelian Randomization Study in the UK Biobank. Arthritis Rheumatol 71, 1634–1641 (2019).

16. Hindy, G. et al. Cardiometabolic Polygenic Risk Scores and Osteoarthritis Outcomes: A Mendelian Randomization Study Using Data From the Malmo Diet and Cancer Study and the UK Biobank. Arthritis Rheumatol 71, 925–934 (2019).

17. Kadam, U.T., Blagojevic, M. & Belcher, J. Statin use and clinical osteoarthritis in the general population: a longitudinal study. J Gen Intern Med 28, 943–9 (2013).

18. Yau, M.S. et al. Validation of knee osteoarthritis case identification algorithms in a large electronic health record database. Osteoarthr Cartil Open 4(2022).

19. Lavin, K.M., Richman, J. S., McDonald, M. N., & Singh, J. A. Osteoarthritis Across Joint Sites in the Million Veteran Program Cohort: Insights From Electronic Health Records and Military Service History. The Journal of rheumatology jrheum.2024-0237(2024).

20. Hunter-Zinck, H. et al. Genotyping Array Design and Data Quality Control in the Million Veteran Program. Am J Hum Genet 106, 535–548 (2020).

21. Honerlaw, J. et al. Framework of the Centralized Interactive Phenomics Resource (CIPHER) standard for electronic health data-based phenomics knowledgebase. J Am Med Inform Assoc 30, 958–964 (2023).

22. L., M.M. Interrater reliability: the kappa statistic. Biochemia medica 22, 276–282 (2012).

23. Westfall, P.H., Troendle, J.F. & Pennello, G. Multiple McNemar tests. Biometrics 66, 1185–91 (2010).

